# Poster and model competition: a novel, inventive, and intriguing teaching-learning method for first-MBBS students

**DOI:** 10.1101/2022.07.18.22277623

**Authors:** Sumera Tasleem, Vaseem Anjum, Syed Imran Ali

**Affiliations:** Department Of Physiology, Malla Reddy Institute Of Medical Sciences,suraram, Hyderabad, Telangana; Department Of Community Medicine, Malla Reddy Institute Of Medical Sciences,suraram, Hyderabad, Telangana; Department Of Physiology, Government Medical College and Hospital, Nagarkurnool

**Keywords:** Poster presentation, Models, Interest, Teaching-learning, Inspiration

## Abstract

**BACKGROUND:** Physiology is one of the foundation subjects of the medical curriculum worldwide. A student cannot grasp the subject thoroughly with didactic lectures alone. As a result, students are developing less interest in learning. Therefore, teachers of Physiology are employing novel, innovative interest-generating learning tools like quizzes, role-plays, seminars, etc besides traditional didactic lectures. Academically poster presentation has been used as a method to resort knowledge. A Poster presentation is a visual treat to the eye and provides an eagle’s view of the topic.

**AIMS AND OBJECTIVES:** A Poster or a Model aims to convey visually and deliver the willful message in an intelligible, crystalline, novel, innovative, interest-generating, unambiguous manner. The study was conducted in the Department of Physiology, Malla Reddy Institute of Medical Sciences for first-year MBBS students. The topic selected was “Cardiovascular Physiology.” The knowledge of the Cardiovascular system is fundamental in understanding crucial concepts, medicine, history-taking of the patient, examination of the CVS in the patient, applied aspects, and pharmacotherapy.

**MATERIALS AND METHODS:** A Poster and Model competition was planned on the same topic and the formal announcement was made regarding the same in routine first-MBBS theory class well in advance, 30 days before the competition day. We divided the class into 14 teams with 14-15 students in each team to ensure a sufficient number in each team if any student dropped. Each team was assigned a different sub-topic for poster and model competition ensuring coverage of the entire portion of cardiovascular physiology. The allotment of the sub-topic was done randomly using the student’s roll numbers. Seven faculty voluntarily agreed to guide students through their poster making, 2 teams were allotted to each faculty by a lottery system to reduce the bias.The study approval was taken from Institutional Ethics Committee. A pre-validated questionnaire was used to take the feedback from both the students and the faculty. The questions were to be answered on a Likert scale from 1 to 5 where 1 strongly disagrees to 5 strongly agree.

**RESULTS:** Out of 200 students,111 participants gave feedback.The male students were 51.4% (n=57) and the female were 48.6% (n=54). The age of the students ranged from 17 to 22 years with a mean of 19.4 ± 1.36 years. Among the faculty, only one male (14.3%) was present, and the rest 85.7% (n=6) were female lecturers. The mean age of the faculty was 36 ± 9.06 years.

The unpaired t-test was applied to all answers and p < 0.05 was taken as significant.Answers was graded as low score(1-2),medium score(3) and high score(4-5).

**CONCLUSION:** The poster/model competition did inculcate enthusiasm in the students. It allowed the students to change their resisting feeling toward the subject which is gratifying for a teacher. This also allowed the faculty to understand the students better. They can identify adverse students by conducting such activities and refine their teaching methods to get better outcomes. Poster/model competition can be used as a novel, innovative interest-generating teaching-learning tool to encourage students and fortify their learning.

## INTRODUCTION

Physiology is one of the foundation subjects of the medical curriculum worldwide. Physiology is the science that seeks to explain the physical and chemical mechanisms that are responsible for the origin, development, and progression of life^1^.A student cannot grasp the subject thoroughly with didactic lectures alone. It is not enough in understanding the fundamental concepts of the subject solo. During the routine lectures, the concentration span can decline after 10-15mins.^2^ As a consequence, students are developing less interest in learning. Therefore, teachers of Physiology are employing novel, innovative interest-generating learning tools like quizzes, role-plays, seminars, etc besides traditional didactic lectures.^3^Academically poster presentation has been used as a method to resort knowledge. A Poster presentation is a visual treat to the eye and provides an eagle’s view of the topic.

A good poster is a summary to deliver knowledge to a student.^4’5^Posters are mind-guide maps to get a better hold of the subject. Poster and model-based learning make use of students’ creative skills. In Model-based learning, students use their creative skills to build models and in the application gain in-depth comprehension and concepts of the concerned topic. After the presentation, students were glad about their achievements and enhanced their assertiveness.3 Students were adaptive to group activity, harmony, and camaraderie during this practice. We make an effort to use posters and models as a teaching-learning tool by conducting a Poster and Model Competition, to seek out how it excites and engenders interest among students concerning a specific topic in physiology.

The study was conducted in the Department of Physiology, Malla Reddy Institute of Medical Sciences for first-year MBBS students. The topic selected was “Cardiovascular Physiology.” The knowledge of the Cardiovascular system is fundamental in understanding crucial concepts, medicine, history-taking of the patient, examination of the CVS in the patient, applied aspects and pharmacotherapy.

## AIMS AND OBJECTIVES

The aim and objective of a Poster or a Model are to convey visually and deliver the willful message in an intelligible, crystalline, novel, innovative, interest-generating, unambiguous manner. In a class of 200 students, a huge changeability in the degree of comprehension, capability, reasoning, inspiration exist. Few students have higher extent of internal inspiration for learning and they will grasp the subject disregarding the teaching method. The traditional teaching method is unlikely to serve the demands of the students who shortfall inbred inspiration. Such students feel declined and lesser inspired to pursue the subject. They find the subject huge, evaporative and burdensome to master and additionally engage; besides finding it tough to comprehend the fundamental postulates of Physiology. We conducted this study for the first-year MBBS students in the embodiment of “posters and model competition” with the ensuing aims and objectives.

1. To ensure an adventitious inspiration for learning and to devolve into internal inspiration during the exercise of poster making and model building.
2. To inculcate profits and ease higher comprehension of the subject and its several concepts so as to help students in their future clinical practice, Physiology being an indispensable part of clinical practice.
3. To incorporate leadership ability to devise and put forward for presentation and induce conviction in them besides furnishing them for future patient conveying and patient comforting.
4. To acknowledge group dynamics of learning and inculcate communion through group exercise, to help students attain workability and flexibility to modify in their group members.Moreover helping students to nuture in consonance and be adjusting in their salute to help them in their future work in life and with others.
5. To tutor students to be at ease in the public, empowering them to control stage fright besides refining their personality and subtle skills.
6. To entitle, educate, and position them to relish studying a topic without making it a difficult affair. This method can be adapted to ease stress among students while studying and make learning pleasurable and educational.
7. To deliver students a chance to explore, unwind and farm their creative expertise, thus combining innovation and knowledge.

## METHODOLOGY

Didactic classroom teaching on the topic of Cardio-vascular systems was taken for the first-MBBS students by the faculty of the department of physiology. A “Poster and Model competition” was planned on the same topic and a formal announcement was made regarding the same in routine first-MBBS theory class well in advance, that is 30 days before the competition day. We divided the class into 14 teams with 14-15 students in each team to ensure a sufficient number in each team if any student dropped. Each team was assigned a different sub-topic for poster and model competition ensuring coverage of the entire portion of cardiovascular physiology. The allotment of the sub-topic was done randomly using the student’s roll numbers. Seven faculty voluntarily agreed to guide students through their poster making and thus 2 teams were allotted to each faculty by a lottery system to reduce the bias.The study was approved by Institutional Ethics Committee, Malla Reddy Institute Of Medical Sciences, Hyderabad.

The following topics were allotted to the students for poster and model presentation

TEAM1:Conducting system of the heart

TEAM2:Microcirculation

TEAM3:Cardiac cycle

TEAM 4:Pathophysiology of shock

TEAM5:Cardiac output

TEAM6:skeletal muscle, cutaneous & splanchic circulation

TEAM7:Blood Pressure

TEAM8:Coronary circulation

TEAM9:Cardiac innervations-Heart rate variability

TEAM10:ECG

TEAM11:Properties of cardiac muscle

TEAM12:Cardiovascular & respiratory changes during exercise

TEAM13:Heart sounds & JVP

TEAM14:Haemodynamics

8. On the day of the competition student presented Poster and Model with great ardour, warmth and vivacity.
9. Feedback forms were given by creating a google form.
10. The best three posters and Models were awarded by the judges.
11. Marks were allotted to each group out of 50 and were included in the internal assessment.
12. The posters and models were evaluated using the following judging criteria
  - Poster introduction/relevance to the theme-10 points
  - Poster explanation/content/presentation-10 points
  - Creativity/originality/attractiveness-10 points
  - Visual impact/overall appearance of poster-10 points
  - Response to question/knowledge/viva-10 points

A pre-validated questionnaire was used to take the feedback from both students and faculty^6’7^. The students’ questionnaire comprised 16 questions on learning feedback, 5 questions were to know if the poster and model competitions are better for assessment, and one regarding interaction feedback. Similarly, the faculty questionnaire had a group of questions to understand the effectiveness of this competition in student learning, assessment, and feedback regarding the mode of learning. The questions were to be answered on a Likert scale from 1 to 5 where 1 strongly disagrees to 5 strongly agree.

## RESULTS

The first MBBS class comprised 200 students. Among them, 111 agreed to give feedback. The unpaired t-test was applied to the questionnaire.Answers were scored based on Likert scale low score(1-2),medium score(4),high score(5).

All 7 faculty members who mentored for the competition were also present on the day of the poster presentation. The male students were 51.4% (n=57) and the female were 48.6% (n=54). The age of the students ranged from 17 to 22 years with a mean of 19.4 ± 1.36 years. The age-gender distribution of the students is depicted in figure 1. Among the faculty, only one male (14.3%) was present, and the rest 85.7% (n=6) were female lecturers. The mean age of the faculty was 36 ± 9.06 years.

**Figure 1:**
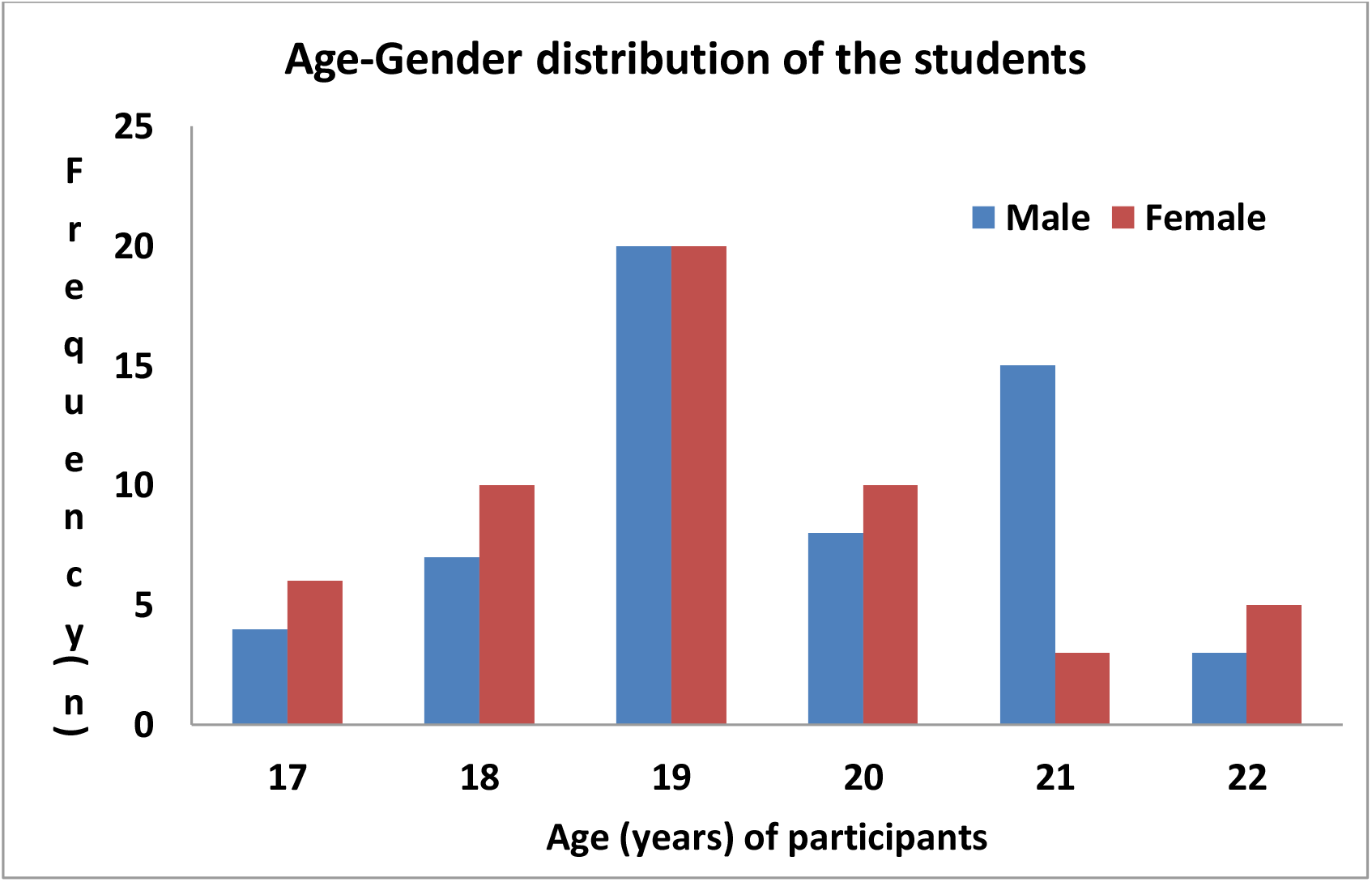
Bar diagram depicting the age-gender distribution in the students (n=111)

The following tables give the details of the response to the questions asked to the students and the faculty.

**Table 1:**
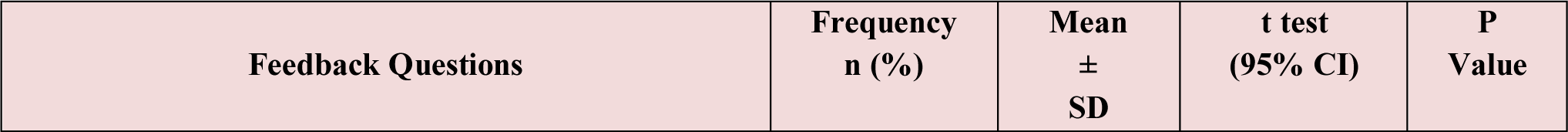
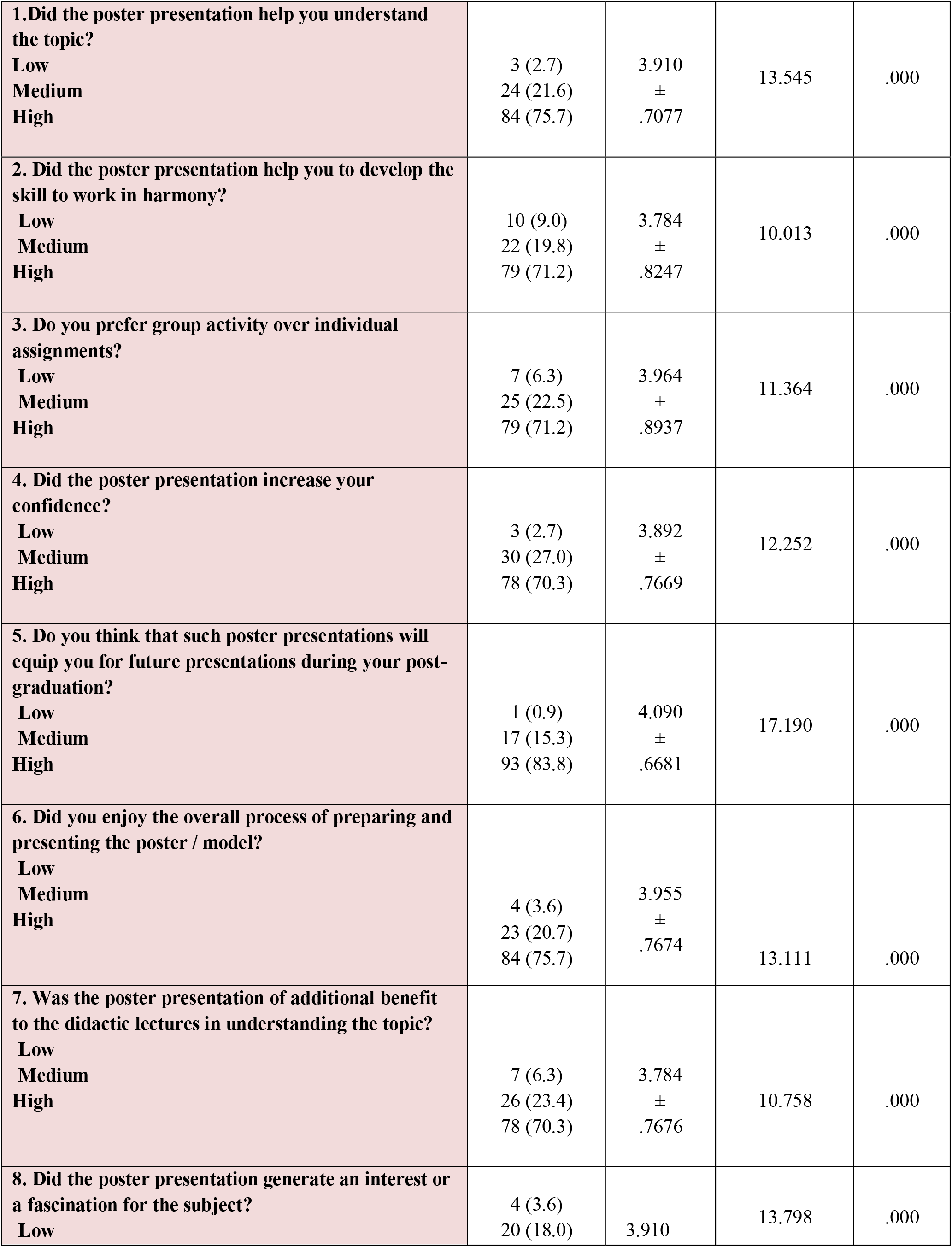
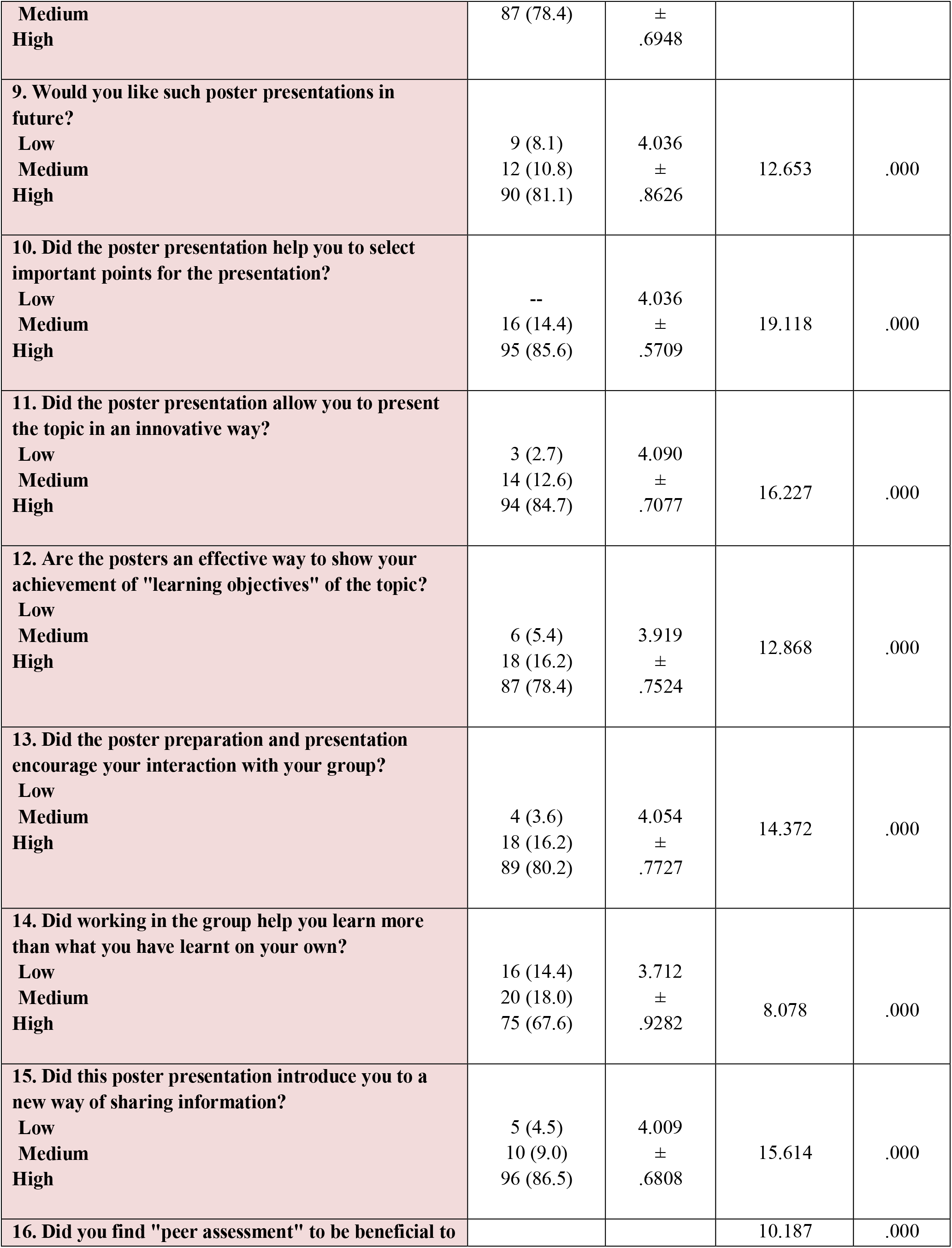
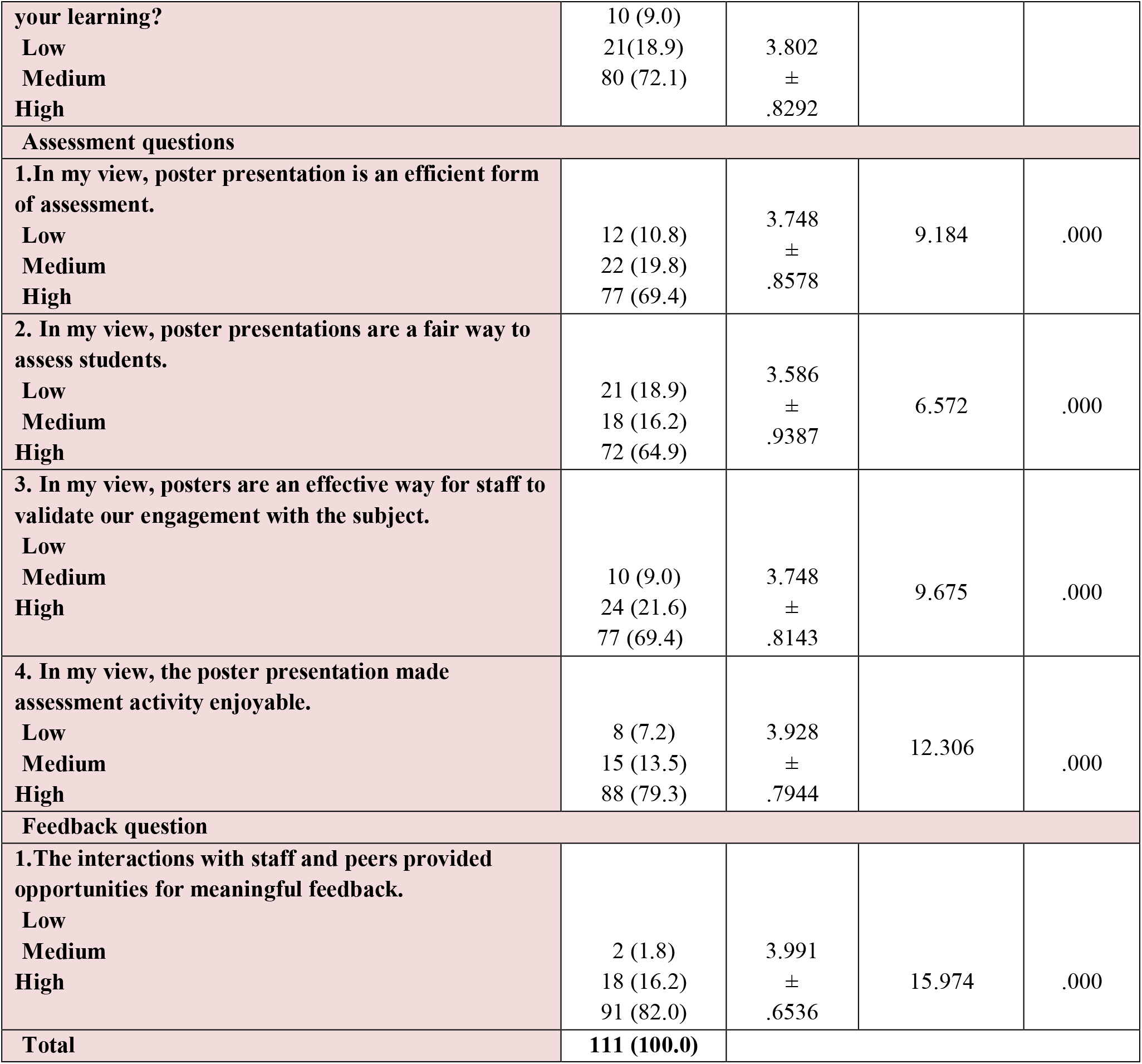
The statistical analysis of the responses of the students (n=111)

**Table 2:**
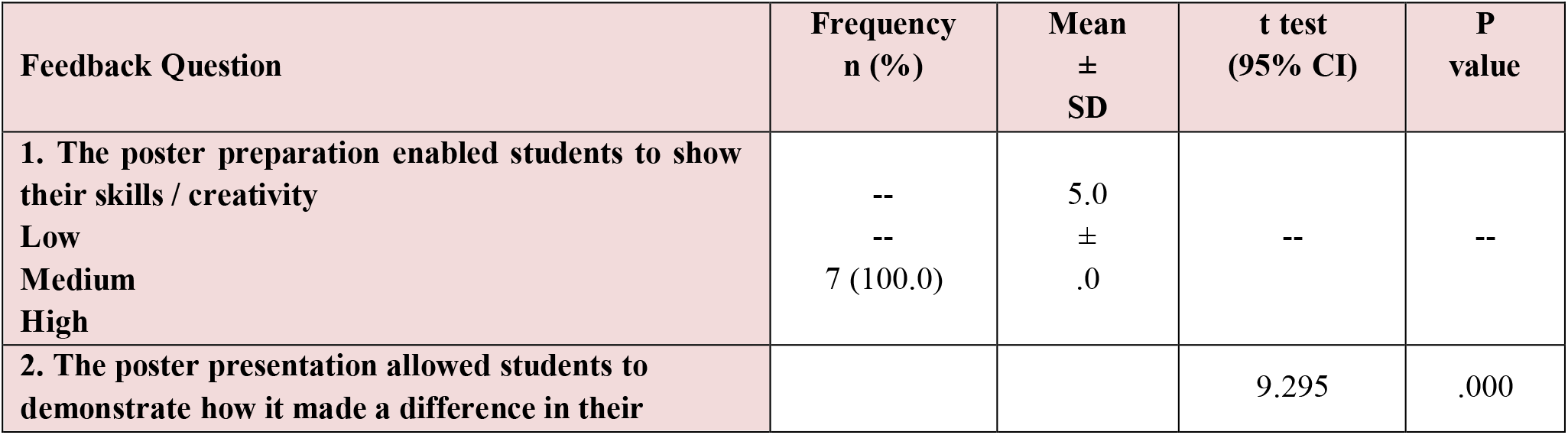
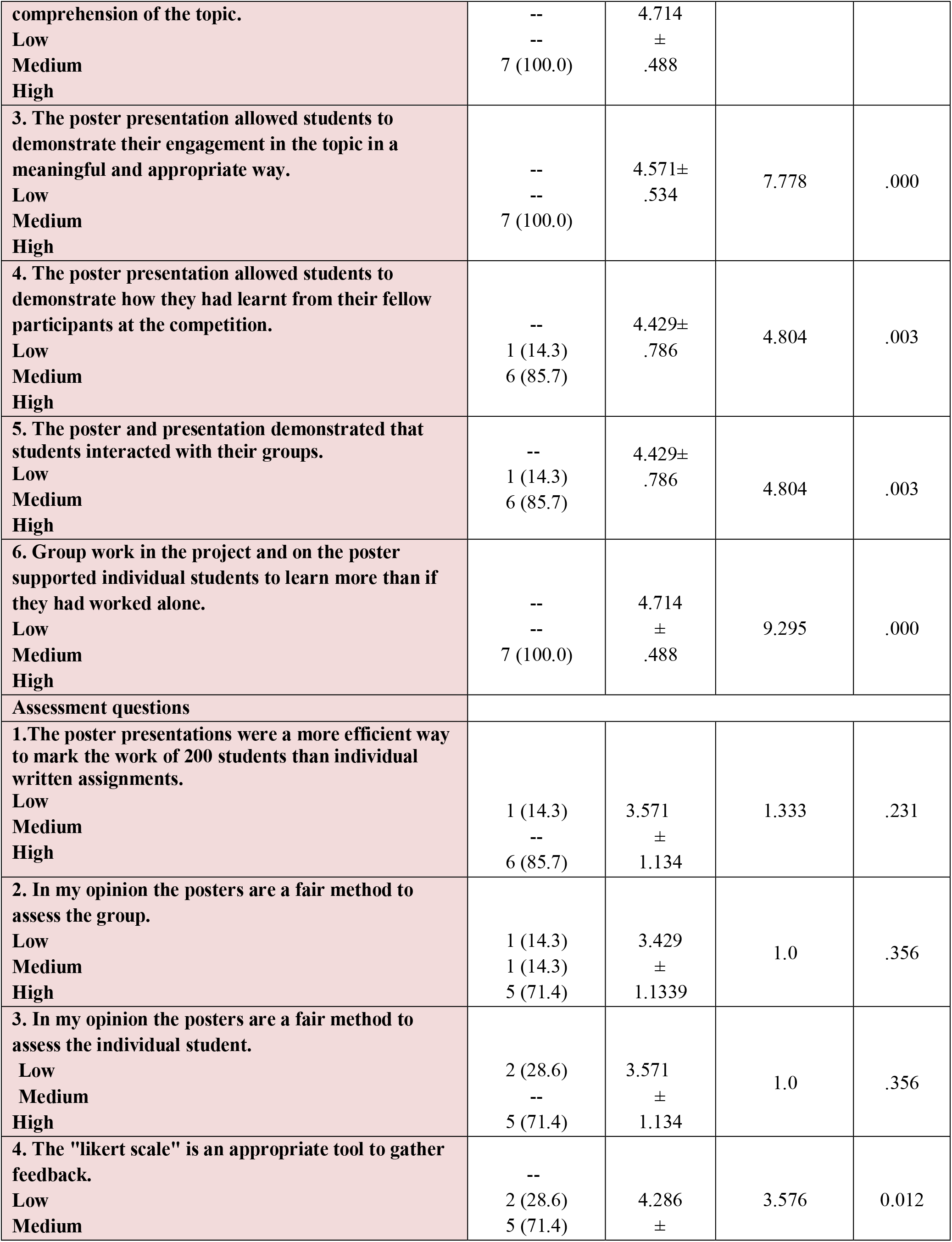
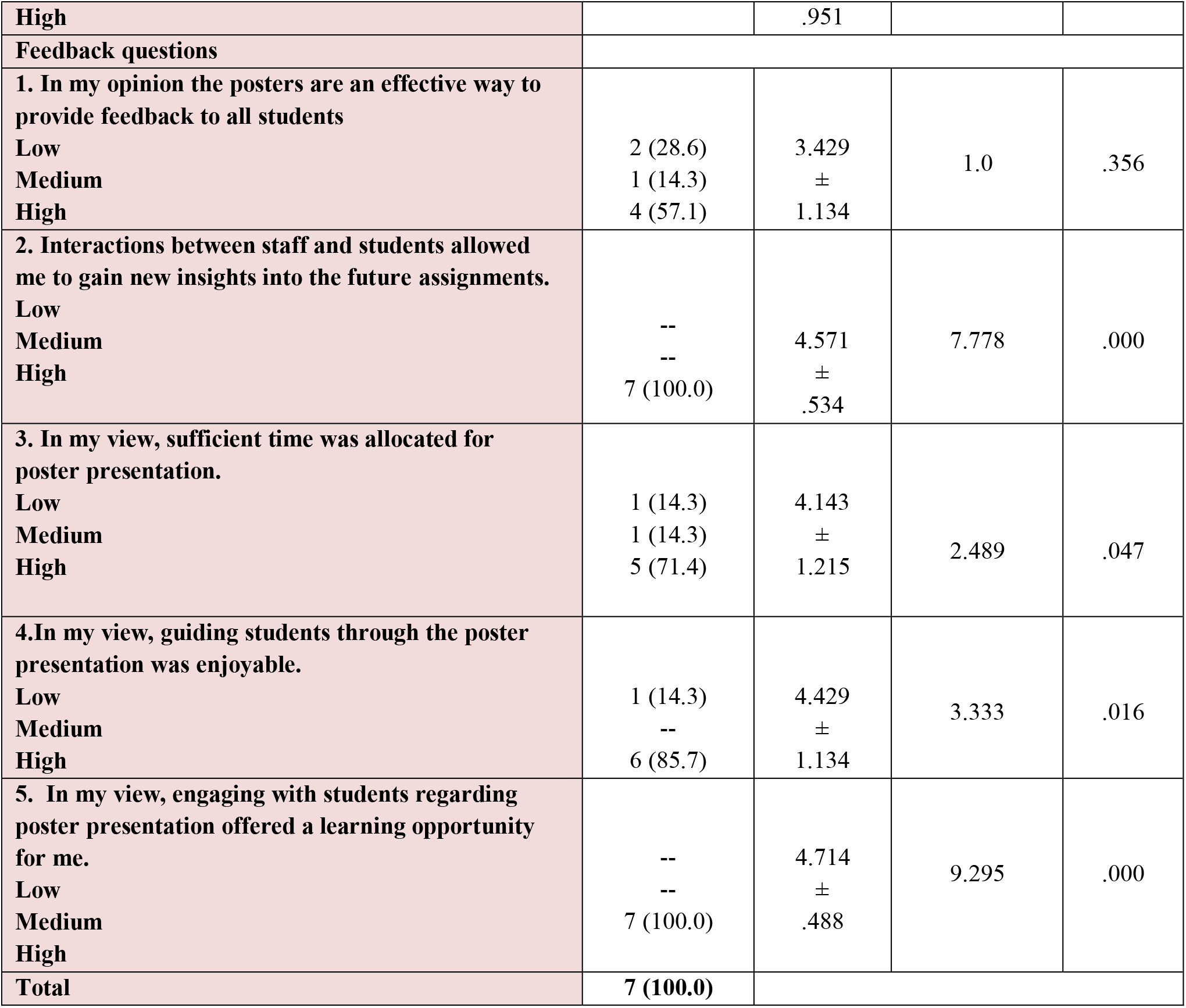
The statistical analysis of the responses of the faculty(n=7)

| **Feedback Question** | **Frequency**<br>**n (%)** | **Mean**<br>**±**<br>**SD** | **t test**<br>**(95% CI)** | **P**<br>**value** |
| --- | --- | --- | --- | --- |
| **1. The poster preparation enabled students to show their skills / creativity**<br>Low<br>Medium<br>High | --<br>--<br>7 (100.0) | 5.0<br>±<br>.0 | -- | -- |
| **2. The poster presentation allowed students to demonstrate how it made a difference in their** |  |  | 9.295 | .000 |
| comprehension of the topic.<br>Low<br>Medium<br>High | --<br>--<br>7 (100.0) | 4.714<br>±<br>.488 |  |  |
| 3. The poster presentation allowed students to demonstrate their engagement in the topic in a meaningful and appropriate way.<br>Low<br>Medium<br>High | --<br>--<br>7 (100.0) | 4.571±<br>.534 | 7.778 | .000 |
| 4. The poster presentation allowed students to demonstrate how they had learnt from their fellow participants at the competition.<br>Low<br>Medium<br>High | --<br>1 (14.3)<br>6 (85.7) | 4.429±<br>.786 | 4.804 | .003 |
| 5. The poster and presentation demonstrated that students interacted with their groups.<br>Low<br>Medium<br>High | --<br>1 (14.3)<br>6 (85.7) | 4.429±<br>.786 | 4.804 | .003 |
| 6. Group work in the project and on the poster supported individual students to learn more than if they had worked alone.<br>Low<br>Medium<br>High | --<br>--<br>7 (100.0) | 4.714<br>±<br>.488 | 9.295 | .000 |
| Assessment questions |  |  |  |  |
| 1.The poster presentations were a more efficient way to mark the work of 200 students than individual written assignments.<br>Low<br>Medium<br>High | 1 (14.3)<br>--<br>6 (85.7) | 3.571<br>±<br>1.134 | 1.333 | .231 |
| 2. In my opinion the posters are a fair method to assess the group.<br>Low<br>Medium<br>High | 1 (14.3)<br>1 (14.3)<br>5 (71.4) | 3.429<br>±<br>1.1339 | 1.0 | .356 |
| 3. In my opinion the posters are a fair method to assess the individual student.<br>Low<br>Medium<br>High | 2 (28.6)<br>--<br>5 (71.4) | 3.571<br>±<br>1.134 | 1.0 | .356 |
| 4. The "likert scale" is an appropriate tool to gather feedback.<br>Low<br>Medium | --<br>2 (28.6)<br>5 (71.4) | 4.286<br>± | 3.576 | 0.012 |
| **High** |  | .951 |  |  |
| **Feedback questions** |  |  |  |  |
| **1. In my opinion the posters are an effective way to provide feedback to all students** |  |  |  |  |
| **Low** | 2 (28.6) | 3.429 | 1.0 | .356 |
| **Medium** | 1 (14.3) | ± |  |  |
| **High** | 4 (57.1) | 1.134 |  |  |
| **2. Interactions between staff and students allowed me to gain new insights into the future assignments.** |  |  |  |  |
| **Low** | -- | 4.571 | 7.778 | .000 |
| **Medium** | -- | ± |  |  |
| **High** | 7 (100.0) | .534 |  |  |
| **3. In my view, sufficient time was allocated for poster presentation.** |  |  |  |  |
| **Low** | 1 (14.3) | 4.143 |  |  |
| **Medium** | 1 (14.3) | ± | 2.489 | .047 |
| **High** | 5 (71.4) | 1.215 |  |  |
| **4. In my view, guiding students through the poster presentation was enjoyable.** |  |  |  |  |
| **Low** | 1 (14.3) | 4.429 | 3.333 | .016 |
| **Medium** | -- | ± |  |  |
| **High** | 6 (85.7) | 1.134 |  |  |
| **5. In my view, engaging with students regarding poster presentation offered a learning opportunity for me.** |  |  |  |  |
| **Low** | -- | 4.714 | 9.295 | .000 |
| **Medium** | -- | ± |  |  |
| **High** | 7 (100.0) | .488 |  |  |
| **Total** | **7 (100.0)** |  |  |  |

## DISCUSSION

It’s difficult for an uninvolved student to accomplish greater comprehension prowess than a dynamic learner^2^. Hence to improve learners’ participation, concerted, related studying would be productive.It is noted that learners who are enterprising in dynamic academic affairs grasp higher than non-compliant learners.^8^The poster/model competition is a creative-based education for laying down external inspiration as many students shortfall innate inspiration. A poster and model competition is an assignment that showcases originality and creativity on the assigned topic after thorough comprehension of the allotted topic.^9^ A project-based assignment is a worthy application to attract students to dynamic learning than traditional didactic lectures.^10^ Always student-focused learning adapts a successful knowledge habitat that enrolls students exclusively in the task.^11^ It delivers different capabilities in a student and motivates them to conceptual learning. The feedback questionnaire analysis takes across opinions postulated by Knowles and others that dynamic learning explains more than static learning.

Analysis of feedback questionnaire

Question 1.Did the poster presentation help you understand the topic?

Many students agreed that exercise helps their comprehension of the topic of cardiovascular physiology. The traditional didactic lectures give a basic notion of the topic, but deeper comprehension was developed by students when they explored and made posters/models on the topic.

It has fortified different concepts previously covered in theory classes. The activity helped them gain additional knowledge on the topic making students apply innovative creativity for presentation. Students created hand-made models and demonstrated them, which enhanced their conceptual learning making everlasting memories.12

Question 2: Did the group activity help you to develop skill to work in harmony?

Many were in agreement with the above interrogation. A systematic, correlative, and reciprocal cluster activity helped nurture esprit among team members and made the poster/model prosperous. This activity made them progressive in their attitude and their essence obliging developing interpersonal skills.This will be beneficial to the students hereafter at their abode.

Question 3. Do you prefer group activity over individual assignments?

71% of the students answered affirmatively. Group activity plows esprit and enhances learners’ knowledge. Interpersonal communication among team members is highly advantageous. Panitz 1996 postulated coordination is a kind of communication for achieving a target objective.^2^ students who did not accomplish well in formative assessment /viva are prone to defective inspiration, subsequently ignoring the chance of learning. For this poster/model activity, they must work in the communion which increases group interaction between dynamic and static learners and helps to overcome academic deficiency among static learners.

Question 4. Did the poster presentation increase your confidence?

Students agreed that it did enhance their confidence, and increased their expertise in the topic of cardiovascular physiology, as it made them do a lot of groundwork which added to their confidence. Justice can be done to any topic if he/she has a deep understanding of the topic, and project it better with mastery and zeal as it involves digging of lot of topics.13

Question 5. Do you think that such poster presentations will equip you for future presentations during your post-graduation?

Ninety-three students(83.8%) answered ‘‘yes’’ to the question. Poster presentation and pedagogy are elements of the post-graduation (PG) timetable. As many students crave PG, this activity will enervate and school them with the principles of preparing a poster/model. The teacher needs to expertise different innovative teaching-learning methods.The poster presentation is fast emerging as a tool for propagating knowledge.^14^ Poster-making is laborious and meticulous.^15^ These skills frame self-assurance in the student, apt for facing difficulties in forthcoming academic tasks.

Question 6: Did you enjoy the process of preparing and presenting the poster/model?

Eighty-four students(75%) agreed the experience was joyful learning. In the words of student wrote “learning can be fun”, joyful experience and thankful to the deparment of Physiology for taking an initiative. Learning by-heart is streneous for many students.In poster/model making they learned different concepts without facing any stress.

Posters are adopted as an approach to disseminating or acquiring knowledge, an innovative tool for studying.^16^ poster presentations develop interpersonal skills with staff and team members.^17’18^

Question 7. Was the poster presentation of additional benefit to the didactic lectures in understanding the topic?

Most of the students (78%) graded it with a high score of 4-5 out of five. In traditional didactic lectures, a basic notion of the topic is provided trying to create interest, but poster/model competition not only fortify traditional lecture but also unwinds surfeit innovative thoughts on the topic for the learner.

Question 8: Did it generate interest or a fascination for the subject?

Intriguingly more than two-thirds (78.4%) found the activity interest generating while only four students said it was not.

This is the undeniable achievement that made us reach our goal that poster/model has inculcated ardor and enhanced knowledge.

Poster/models approached learning physiology as scenic, fascinating and alluring. Model making is charismatic. It can bring out the creative side in all of us. A perfect model can bewitch the passion of students and ineradicable emboss on their brains. The scenery will punch into their brain easily.^19^

This activity showcases their talent, originality, and creativity in a different innovative way.^20^

Question 9: Would you like such presentation in the future?

The majority of the students(81%) agreed with the query, which motivated us and reassured us that learning can be enjoyable. The poster presentation can be adopted as a teaching-learning method for students.^22^

Peer-to-peer presentations to fellow batch mates and on an official platform yield discourse on a broader aspect.^16^ some motivation from the lecturers can prowess their originality as all are adult learners.^21^

Question 10. Did the poster presentation help you to select important points for the presentation?

Ninety-five(85%) agreed to this query. In order to reach this target students referred multiple sources for the topic in detail and made poster/model.Students developedfeeling of procurement when they prepared and presented the poster/model, thus making them adapt to various learning modes.The entire activity kindled attentiveness in the topic.

Question 11. Did the poster presentation allow you to present the topic in an innovative way? Many of the students (84.7%) said yes to the query.Many students felt poster way better then one –way teaching as the activity required leadership, peer interaction, skills, co-ordination.Students liked the whole concept of presenting a topic in the form of poster/model and they felt they have mastered the topic.The activity increased their confidence in the topic.few students said in their words’ we want posters in every topic’,whereas other said ‘we want to make posters in difficult topics like central nervous system’.This exercise was a enjoyable task as opposed to vivas and tests.^17^

Question 12:Are the posters an effective way to show your achievement of “learning objectives” of the topic?

Eighty-seven(78.4%) said yes to the query. Students were given learning objectives before preparing a poster/model accordingly. External inspiration by the lecturer accelerated their internal inspiration which was self-perpetuating.23The student underwent an intellectual variation during the exercise. This gave them a sense of achievement and made learning pleasurable.

Question 13:Did the poster preparation and presentation encourage your interaction with your group?

Eighty-nine(80.2%) agreed to the query. For students who were perverted, and alienated, this activity developed their communication skills and self-confidence. The journey of poster-making was energetic and joyful and in the course, students made new friends, shared knowledge, and cleared their doubts.

Question 14:Did working in the group help you learn more than what you have learnt on your own?

Students felt poster/model created occasion for intellectual learning, student-student, and student-faculty interaction made a bond with them, built good memories of the MBBS course which they can preserve life-long.^14^

Question 15. Did this poster presentation introduce you to a new way of sharing information? Many students(86.5%) appreciated the notion of the poster presentation. They want more of it in the future a student commented. Few found this activity deviation from routine lecture class. Students interacted much with faculty and in the process mastered different artistic skills. A poster can be used as an assessment tool for their innovative thoughts, and creative skills. Students could complete their tasks in a tension-free environment.

Question 16. Did you find “peer assessment” to be beneficial to your learning?

A majority (72.1%) agreed as students with communication skills developed interpersonal skills and motivation. Students who lost to learning reinforced during the exercise, such students may have avoided learning for the entire year.Teachers are the driving force for students to attain cavernous learning.^24^

Many of the Staff members felt positive during the poster/model competition. Posters/models enabled students to show their creative skills, and originality made difference in their cognition of the topic, and made student-staff interaction substantial which aided maximum learning during and after the exercise. Difficulties and doubts during individual learning were subsided by peer-peer interaction, accelerated communication skills, enhanced knowledge of the individual, and learned mutual group harmony. Posters are a useful learning tool. Posters/models made the topic pleasurable. It summarized the entire topic in one go. Students were satisfied with the assessment criteria. Students were judged on knowledge of the topic, presentation, originality, creativity, demonstration, group skills, confidence, and communication skills. This will also help them in future academic endeavors.

Teachers should be role models.^21^ A teacher should not only take a class but entertain novel, innovative, academic activities to develop an interest in learning. A teacher should always motivate students in adopting novel tools of learning.

## CONCLUSION

A single method of teaching is not enough. Gaining knowledge should be pleasurable. It was delightful and visual-treat to see students present posters/models with a lot of ardor and zeal. Students changed their likes and dislikes during their exercise, which is rewarding for a teacher. A teacher must identify adverse students and refine their learning by conducting novel activities to get better outcomes. The poster/ model competition can be used as a method of learning as it is interest-generating and encourage students to gain knowledge in a pleasurable way.

Poster/model competition can be used as a novel, innovative interest-generating teaching-learning tool to encourage students and fortify their learning.

It also allowed the students to change their resisting feeling toward the subject which is gratifying for a teacher. This also allowed the faculty to understand the students better. They can identify adverse students by conducting such activities and refine their teaching methods to get better outcomes.

## Data Availability

All data produced in the present study are available upon reasonable request to the authors
All data produced in the present work are contained in the manuscript

## Ethical approval

The study was approved by the Institutional Ethics Committee

## REFERENCES

1. Guyton and Hall. Textbook of medical physiology. Third south Asia Edition.chapter 1.pg no3. New Delhi:Elsevier publication 2021.ISBN:978-0-323-59712-8

2. Murray N. Improving the way technical skills are taught in creative studio-an exploration of student centered learning method. Transaction. 2012;9(2):30–58.

3. Moule P, Judd M, Girot E. The poster presentation: What value to the teaching and assessment of research in pre- and post-registration nursing courses? Nurse Educ Today. 1998;18(3):237–42.

4. Moneyham L, Ura D, Ellwood S, Bruno B. The poster presentation as an educational tool. Nurse Educ. 1996;21(4):45–7.

5. Rowe N, Ilic D. What impact do posters have on academic knowledge transfer? A pilot survey on author attitudes and experiences. BMC Med Educ. 2009;9:71.

6. Lois James Samuel, Laveena V. Bandodkar, Padmanabh V. Rataboli. Poster and model competition: a novel interest-generating teaching tool in the subject of pharmacology. International Journal of Basic & Clinical Pharmacology. doi: 10.5455/2319-2003.ijbcp20140816.

7. Ross et al. BMC Medical Education (2019) 19:432. Using poster presentation to assess large classes: a case study of a first-year undergraduate module at a South African university. https://doi.org/10.1186/s12909-019-1863-9.

8. Hiemstra R, Sisco B. Individualising instruction. Moving from Pedagogy to Androgogy. San Francisco: JosseyBass; 1990. Available from: http://www.distance.syr.edu/androgogy.html. [Last accessed on 2014 Mar 20].

9. Graham R, Crawley E. Making Projects Work: a review of transferable best practice approaches to engineering. Project based learning in UK. Engineering Education. The Higher Education Academy Journal, 5(2):41–9. Available from: http://www.journals.headacademy.ac.uk/doi/full/10.11120/ened:2010.

10. Blumenfeld PC, Soloway E, Ronald W, Krajcik JS, Guzdial M, Palincsar A. Motivating project based learning: sustaining, supporting the learning. Educ Psychol. 1991;26(3-4):369–98.

11. Bradford M. Motivating students through project based service learning. THE J 2005;32(6):29. ERIC 6.

12. Crooks DL, Kilpatrick M. In the eye of the beholder: Making the most of poster presentations – Part 2. Can OncolNurs J 1998;8:154–9.

13. Rowe N, Ilic D. Poster presentation - A visual medium for academic and scientific meetings. PaediatrRespir Rev. 2011;12(3):208–13.

14. Moore LW, Augspurger P, King MO, Proffitt C. Insights on the poster preparation and presentation process. ApplNurs Res. 2001;14(2):100–4.

15. Hardicre J, Coad J, Devitt P. Ten steps to successful conference presentations. Br J Nurs. 2007;16(7):402–4.

16. Pelletier D. The focused use of posters for graduate education in the complex technological nursing environment. Nurse Educ Today. 1993;13(5):382–8.

17. Boullata JI, Mancuso CE. A “how-to” guide in preparing abstracts and poster presentations. NutrClinPract. 2007;22(6):641–6.

18. Taggart HM, Arslanian C. Creating an effective poster presentation. OrthopNurs. 2000;19(3):47-9, 52.

19. Brodsky L. In: Whaler PG, editor. Why model? Professional model making, history, practice, tips, tricks and thinking loud. Saturday Aug 21, 2010. Available from: http://www.promodelmaking.wordpress.com. [Last accessed on 2013 Nov 11].

20. Chaffee H. Ten talents of a model maker. Available from: http://www.Modelmaker.org, http://www.serc.carleton.edu/introgeo/models/. [Last accessed on 2013 Apr 22].

21. Bhuiyan PS, Rege NN, Supe ON. Learning and motivation. In: Parkar SR, Shah F, editor. The Art of Teaching Medical Students.Chapter 6. Maharashtra: Medical Education Technology Cell, Seth G.S. Medical College and KEM Hospital.

22. Accessed from: www.bie.org/PBLstrategies. accessed on Nov 28, 2013.

23. Thomas M. Effective Teaching.Chapter 2. Motivation an energizer. Ramnagar, Delhi: S. Chand and Co.; 2008: 20–3.

24. Hardie M. An inquiry based learning approach to teaching about planning regulations. Transactions. 2009;6(2):5–18. [Last accessed on 2014 Feb 05].

