## supplemental Table for "Poster and model competition: a novel, inventive, and intriguing teaching-learning method for first-MBBS students": poster tables.docx

**Table 1: The statistical analysis of the responses of the students (n=111)**

| **Feedback Questions** | **Frequency**  **n (%)** | **Mean**  **±**  **SD** | **t test**  **(95% CI)** | **P**  **Value** |
| --- | --- | --- | --- | --- |
| **1.Did the poster presentation help you understand the topic?**  **Low**  **Medium**  **High** | 3 (2.7)  24 (21.6)  84 (75.7) | 3.910  ±  .7077 | 13.545 | .000 |
| **2. Did the poster presentation help you to develop the skill to work in harmony?**  **Low**  **Medium**  **High** | 10 (9.0)  22 (19.8)  79 (71.2) | 3.784  ±  .8247 | 10.013 | .000 |
| **3. Do you prefer group activity over individual assignments?**  **Low**  **Medium**  **High** | 7 (6.3)  25 (22.5)  79 (71.2) | 3.964  ±  .8937 | 11.364 | .000 |
| **4. Did the poster presentation increase your confidence?**  **Low**  **Medium**  **High** | 3 (2.7)  30 (27.0)  78 (70.3) | 3.892  ±  .7669 | 12.252 | .000 |
| **5. Do you think that such poster presentations will equip you for future presentations during your post-graduation?**  **Low**  **Medium**  **High** | 1 (0.9)  17 (15.3)  93 (83.8) | 4.090  ±  .6681 | 17.190 | .000 |
| **6. Did you enjoy the overall process of preparing and presenting the poster / model?**  **Low**  **Medium**  **High** | 4 (3.6)  23 (20.7)  84 (75.7) | 3.955  ±  .7674 | 13.111 | .000 |
| **7. Was the poster presentation of additional benefit to the didactic lectures in understanding the topic?**  **Low**  **Medium**  **High** | 7 (6.3)  26 (23.4)  78 (70.3) | 3.784  ±  .7676 | 10.758 | .000 |
| **8. Did the poster presentation generate an interest or a fascination for the subject?**  **Low**  **Medium**  **High** | 4 (3.6)  20 (18.0)  87 (78.4) | 3.910  ±  .6948 | 13.798 | .000 |
| **9. Would you like such poster presentations in future?**  **Low**  **Medium**  **High** | 9 (8.1)  12 (10.8)  90 (81.1) | 4.036  ±  .8626 | 12.653 | .000 |
| **10. Did the poster presentation help you to select important points for the presentation?**  **Low**  **Medium**  **High** | --  16 (14.4)  95 (85.6) | 4.036  ±  .5709 | 19.118 | .000 |
| **11. Did the poster presentation allow you to present the topic in an innovative way?**  **Low**  **Medium**  **High** | 3 (2.7)  14 (12.6)  94 (84.7) | 4.090  ±  .7077 | 16.227 | .000 |
| **12. Are the posters an effective way to show your achievement of "learning objectives" of the topic?**  **Low**  **Medium**  **High** | 6 (5.4)  18 (16.2)  87 (78.4) | 3.919  ±  .7524 | 12.868 | .000 |
| **13. Did the poster preparation and presentation encourage your interaction with your group?**  **Low**  **Medium**  **High** | 4 (3.6)  18 (16.2)  89 (80.2) | 4.054  ±  .7727 | 14.372 | .000 |
| **14. Did working in the group help you learn more than what you have learnt on your own?**  **Low**  **Medium**  **High** | 16 (14.4)  20 (18.0)  75 (67.6) | 3.712  ±  .9282 | 8.078 | .000 |
| **15. Did this poster presentation introduce you to a new way of sharing information?**  **Low**  **Medium**  **High** | 5 (4.5)  10 (9.0)  96 (86.5) | 4.009  ±  .6808 | 15.614 | .000 |
| **16. Did you find "peer assessment" to be beneficial to your learning?**  **Low**  **Medium**  **High** | 10 (9.0)  21(18.9)  80 (72.1) | 3.802  ±  .8292 | 10.187 | .000 |
| **Assessment questions** | | | | |
| **1.In my view, poster presentation is an efficient form of assessment.**  **Low**  **Medium**  **High** | 12 (10.8)  22 (19.8)  77 (69.4) | 3.748  ±  .8578 | 9.184 | .000 |
| **2. In my view, poster presentations are a fair way to assess students.**  **Low**  **Medium**  **High** | 21 (18.9)  18 (16.2)  72 (64.9) | 3.586  ±  .9387 | 6.572 | .000 |
| **3. In my view, posters are an effective way for staff to validate our engagement with the subject.**  **Low**  **Medium**  **High** | 10 (9.0)  24 (21.6)  77 (69.4) | 3.748  ±  .8143 | 9.675 | .000 |
| **4. In my view, the poster presentation made assessment activity enjoyable.**  **Low**  **Medium**  **High** | 8 (7.2)  15 (13.5)  88 (79.3) | 3.928  ±  .7944 | 12.306 | .000 |
| **Feedback question** | | | | |
| **1.The interactions with staff and peers provided opportunities for meaningful feedback.**  **Low**  **Medium**  **High** | 2 (1.8)  18 (16.2)  91 (82.0) | 3.991  ±  .6536 | 15.974 | .000 |
| **Total** | **111 (100.0)** |  | | |

**Table 2: The statistical analysis of the responses of the faculty(n=7)**

| **Feedback Question** | **Frequency**  **n (%)** | **Mean**  **±**  **SD** | **t test**  **(95% CI)** | **P**  **value** |
| --- | --- | --- | --- | --- |
| **1. The poster preparation enabled students to show their skills / creativity**  **Low**  **Medium**  **High** | --  --  7 (100.0) | 5.0  ±  .0 | -- | -- |
| **2. The poster presentation allowed students to demonstrate how it made a difference in their comprehension of the topic.**  **Low**  **Medium**  **High** | --  --  7 (100.0) | 4.714  ±  .488 | 9.295 | .000 |
| **3. The poster presentation allowed students to demonstrate their engagement in the topic in a meaningful and appropriate way.**  **Low**  **Medium**  **High** | --  --  7 (100.0) | 4.571±  .534 | 7.778 | .000 |
| **4. The poster presentation allowed students to demonstrate how they had learnt from their fellow participants at the competition.**  **Low**  **Medium**  **High** | --  1 (14.3)  6 (85.7) | 4.429±  .786 | 4.804 | .003 |
| **5. The poster and presentation demonstrated that students interacted with their groups.**  **Low**  **Medium**  **High** | --  1 (14.3)  6 (85.7) | 4.429±  .786 | 4.804 | .003 |
| **6. Group work in the project and on the poster supported individual students to learn more than if they had worked alone.**  **Low**  **Medium**  **High** | --  --  7 (100.0) | 4.714  ±  .488 | 9.295 | .000 |
| **Assessment questions** |  | | | |
| **1.The poster presentations were a more efficient way to mark the work of 200 students than individual written assignments.**  **Low**  **Medium**  **High** | 1 (14.3)  --  6 (85.7) | 3.571  ±  1.134 | 1.333 | .231 |
| **2. In my opinion the posters are a fair method to assess the group.**  **Low**  **Medium**  **High** | 1 (14.3)  1 (14.3)  5 (71.4) | 3.429  ±  1.1339 | 1.0 | .356 |
| **3. In my opinion the posters are a fair method to assess the individual student.**  **Low**  **Medium**  **High** | 2 (28.6)  --  5 (71.4) | 3.571  ±  1.134 | 1.0 | .356 |
| **4. The "likert scale" is an appropriate tool to gather feedback.**  **Low**  **Medium**  **High** | --  2 (28.6)  5 (71.4) | 4.286  ±  .951 | 3.576 | 0.012 |
| **Feedback questions** |  | | | |
| **1. In my opinion the posters are an effective way to provide feedback to all students**  **Low**  **Medium**  **High** | 2 (28.6)  1 (14.3)  4 (57.1) | 3.429  ±  1.134 | 1.0 | .356 |
| **2. Interactions between staff and students allowed me to gain new insights into the future assignments.**  **Low**  **Medium**  **High** | --  --  7 (100.0) | 4.571  ±  .534 | 7.778 | .000 |
| **3. In my view, sufficient time was allocated for poster presentation.**  **Low**  **Medium**  **High** | 1 (14.3)  1 (14.3)  5 (71.4) | 4.143  ±  1.215 | 2.489 | .047 |
| **4.In my view, guiding students through the poster presentation was enjoyable.**  **Low**  **Medium**  **High** | 1 (14.3)  --  6 (85.7) | 4.429  ±  1.134 | 3.333 | .016 |
| **5. In my view, engaging with students regarding poster presentation offered a learning opportunity for me.**  **Low**  **Medium**  **High** | --  --  7 (100.0) | 4.714  ±  .488 | 9.295 | .000 |
| **Total** | **7 (100.0)** |  | | |
